# Communication of pain intensity and unpleasantness through magnitude ratings: Influence of scale type, but not sex of the participant

**DOI:** 10.1101/2021.06.24.21259433

**Authors:** Hadas Nahman-Averbuch, Cassidy Hughes, Marie-Eve Hoeppli, Kristina White, James Peugh, Eric Leon, Christopher D. King, Robert C. Coghill

## Abstract

A critical aspect for much human pain research is the ability of participants to communicate their first person, experiential perspective to a third person observer. This communication is frequently accomplished via pain ratings. The type of scale and how participants/patients may differentially use the scale has a major influence on the communication of pain experiences. The present study examined the role of sex on the pain rating process using both noxious and innocuous stimuli and two different types of rating scales. Participants underwent noxious heat, auditory and visual stimulation paradigms. Pain intensity and unpleasantness ratings were collected using the visual analog scale (VAS) and numerical rating scale (NRS) in a random order. For noxious heat stimuli, low (44-45°C) and high (47-48°C) intensity stimuli were delivered. To identify if one rating scale allows better discrimination between different stimulus intensities and if this is dependent on sex, discrimination thresholds were calculated. Significant effect for rating scale and intensity level of stimuli were found for all stimulus modalities (noxious heat, auditory and visual) indicating that higher intensity and unpleasantness ratings were found using the NRS compared to the VAS. No effect of sex or interaction with sex was found. No differences in rating scale and sex were found for the discrimination thresholds. Biases in rating scales usage exist with NRS yielding higher ratings to the same stimuli. However, this bias does not appear to contribute significantly to sex differences in pain.

## 1. Introduction

Human pain research, clinical trials and pain management often rely on the ability of participants or patients to communicate their experience. This communication is frequently accomplished via pain ratings [18]. Many factors can affect the communication of pain experiences including the type of rating scale and the participants/patients usage of the scale. Two of the most commonly used pain scales to quantify pain sensation are the NRS and VAS [10, 26]. Both the NRS and the VAS have been validated for ratings of pain magnitude [5, 8, 9, 20] and ratings on both scales are highly correlated in healthy participants and pain patients [2, 5, 8, 10, 11, 14, 26]. However, higher ratings are often found with NRS [3, 11, 19], which may be due to key differences between the scales. The VAS, but not the NRS, has ratio scale properties [20]. In addition, patients appear to use the NRS as a categorical scale while they use the VAS as a continuous scale [6] [12]. For example, for the NRS scale, participants are more likely to use integers, while for the VAS, participants are more likely to use the full range of the scale [6] [12]. This different usage of scales may also impact pain discrimination by altering the ability to report small differences in pain intensity. A previous study found that the participants using the VAS can better discriminate between pairs of adjacent heat stimuli (difference of 1°C) compared to when they use the verbal descriptor scale [24].

Another factor that can influence the communication of pain experiences and pain ratings is sex. Contradicting evidence for sex differences in experimental pain responses in healthy participants exists. Some studies report that healthy females have higher pain sensitivity compared to healthy males while others do not [4, 15, 22, 23]. The different findings may be related to the stimulus modality, measure, and location [4, 23]. Another potential factor is that males and females may report and communicate pain differently because of different usage of rating scales [1, 7, 8, 13, 21]. In healthy participants, sex differences for cold pain ratings have been found when using the numerical rating scale (NRS) but not the visual analog scale (VAS) [8]. In addition, sex differences have been found for heat stimuli when using an electronic NRS but not a VAS scale [21].

The present study examined the role of sex on the pain rating process using both noxious and innocuous stimuli and two different types of rating scales (NRS vs. VAS). The use of various modalities (noxious heat, auditory, and visual stimuli) allowed us to differentiate between general biases in scale usage and biases that are specific to pain. The primary hypothesis was that sex differences would be found when using NRS but not VAS. The secondary hypothesis was that greater discrimination would be found with the ratings obtained with the VAS compared to those obtained with the NRS.

## 2. Methods

This study was approved by the Institutional Review Board of Cincinnati Children’s Hospital Medical Center. Written, informed consent was obtained from all participants prior to participating in the study. In case of participants that are < 18 years old, written, informed consent was obtained from participants’ parent or guardian and assent was obtained from all participants prior to participating in the study.

### 2.1. Study Participants

Inclusion criteria for all study participants were: (1) males or females, (2) ages 14-44, (3) generally healthy. Exclusion criteria were: (1) having chronic pain, (2) diabetes, (3) skin conditions on the forearms (4) neurological disorder, (5) taking medications for pain or migraine, (6) taking medications for neurological or psychiatric disorders, (7) use of a prescription or recreation drug which affects the central nervous system (tested by a urine drug screen at the beginning of the study). In addition, all participants were naïve and did not previously participate in a psychophysical study of pain involving either the VAS or the NRS.

### 2.2. Study design

All procedures were completed in a single visit. Participants first completed the familiarization portion, then they completed the noxious heat, auditory and visual paradigms. There were two modality orders, and within each order, scale usage was randomized (Fig. 1). Half-way through the testing portion, participants completed self-report psychological surveys for anxiety, depression, pain catastrophizing, and neuroticism to characterize the sample population and examine potential sex differences of these measures. The study visit was completed in about two hours.

**Figure 1.**
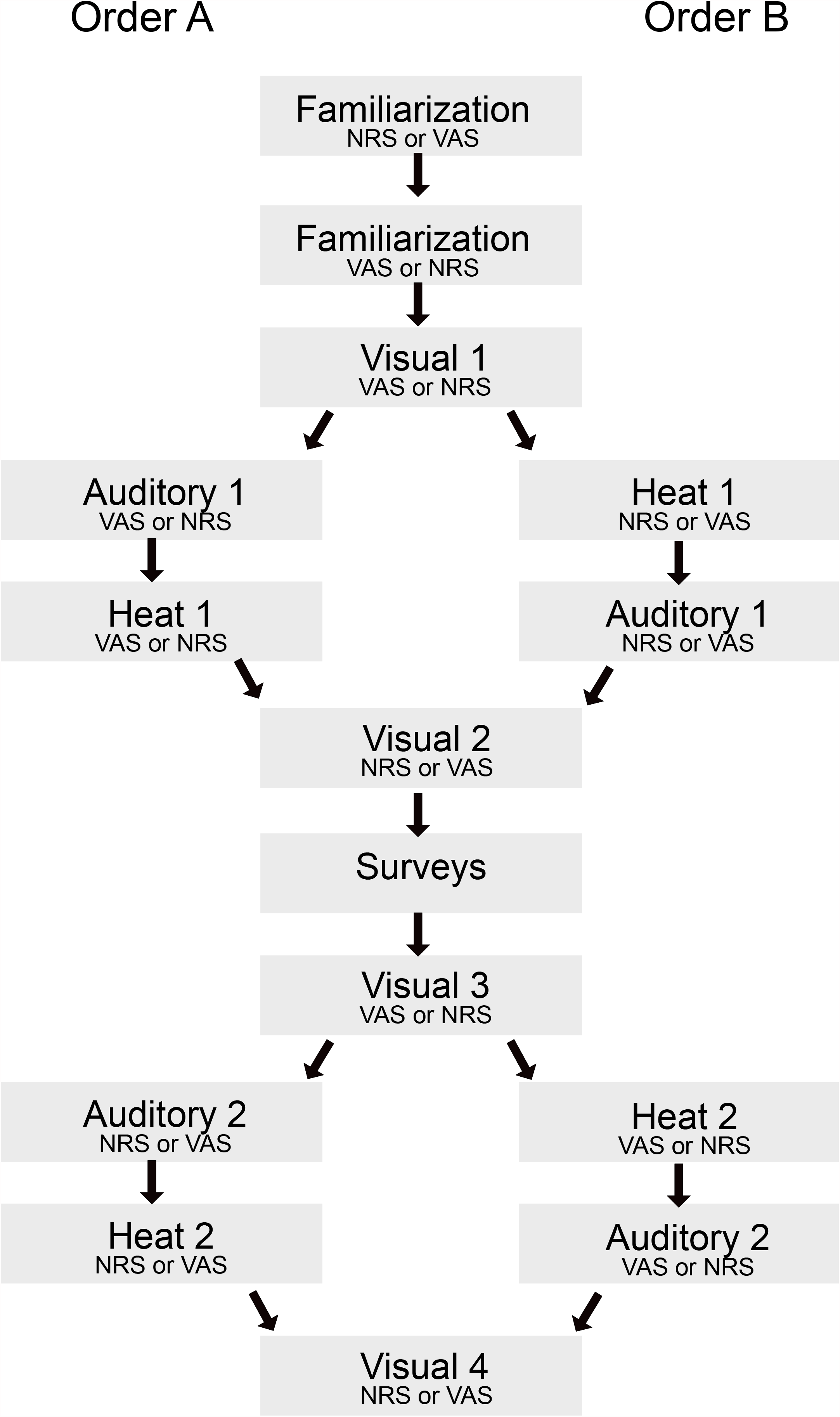
Study design. **A**. After a familiarization phase (conducted separately for NRS and VAS), participants completed heat, auditory, and visual paradigms in a random order. Participants rated each stimulus using both an NRS and a VAS, but the choice of scale was randomized for each paradigm. The order of stimuli within each paradigm was also randomized. **B**. In the heat paradigm, high and low intensity stimuli were presented once in a random order. In the auditory paradigm, stimuli were presented twice. For the visual paradigm, stimuli were presented once.

### 2.3. Rating Scales

Participants were asked to rate various heat, auditory, and visual stimuli with a VAS and an NRS. Only one scale type was used for a given block of stimuli. The order of scale usage for all stimuli modalities was randomized for each participant. Pain intensity and pain unpleasantness ratings for heat stimuli were collected separately, as were ratings of sound loudness and unpleasantness for auditory stimuli. For visual stimuli, only intensity ratings were collected.

Pain intensity (how strong the pain feels) and pain unpleasantness (how disturbing the pain feels) were defined using a radio analogy [20]. As the scales were described to the participants, the same anchors of no pain/unpleasantness to the most intense pain/unpleasantness imaginable were used for both the VAS and the NRS rating scales.

#### VAS

A mechanical VAS was used in this study. Participants were able to slide the inner portion of the VAS. The further they slide to the right with more color exposed (on the participant facing side) corresponded to greater pain intensity or unpleasantness. Ratings corresponded to numeric values 0-10 accurate to one decimal place (on the experimenter-facing side). Participants were not allowed to see the numeric values of their VAS ratings.

#### NRS

Participants were asked to rate the pain verbally from 0 (no pain/unpleasantness)-10 (the most intense pain/unpleasantness imaginable). Decimals were accepted if the participant chose to use them but were not explicitly presented as an option.

### 2.4. Sensory Testing

#### 2.4.1. Familiarization

Each participant engaged in a stimulus-response heat paradigm to familiarize them with the heat stimuli and rating scales. The paradigm involved 16 heat stimuli ranging from 35-49°C (35, 43, 44, 45, 46, 47, 48, 49°C, 5 second plateau duration, rise/fall rates 6°C/s) delivered to the left forearm using a 16 × 16 mm thermode (Medoc, Ramat Yishai, Israel). For the first 8 stimuli, the temperatures were presented in ascending order, and in the last 8 stimuli, they were presented in a pseudo-randomized order. Baseline temperature was 35°C, and there was a thirty second break between stimuli. Immediately after the stimulus temperature returned to baseline, the participant was asked to rate the pain intensity and pain unpleasantness from the stimulus successively. The stimulus-response heat paradigm was presented twice, once using the VAS and once the NRS. Scale order was pre-randomized for each participant.

#### 2.4.2. Test Heat Stimuli

Two heat paradigms were delivered. Each paradigm included 10 stimuli delivered to the left forearm. Each heat stimulus was five-second plateau duration (rise/fall 6°C/s), baseline temperature was 35°C with fifteen seconds between each stimulus (16 × 16 mm thermode; Medoc, Ramat Yishai, Israel). Each paradigm included lower intensity stimuli (44, 44.2, 44.5, 44.7, 45°C) and higher intensity stimuli (47, 47.2, 47.5, 47.7, 48°C) in a randomized order (high intensities first or low intensities first). In addition, stimuli within the low or the high intensities were pseudorandomized with two sequences exist for each intensity (2 for low intensity and 2 for high intensity). Each participant was pre-assigned to complete two paradigms, one for each scale (VAS and NRS). Order of scale usage was also randomized between participants. Participants were asked to rate both pain intensity and pain unpleasantness after each stimulus.

#### 2.4.3. Auditory Stimuli

There were two different auditory paradigms. Each paradigm had eight total stimuli with 4 different stimulus intensity levels: 68, 71, 75, and 80 decibels. Each auditory stimulus had a plateau duration of five seconds (rise/fall rates 41 dB/s), and there a was a fifteen second break (silence) between each stimulus. The paradigms were pseudo-randomized such that all intensity levels were presented once before any intensity was repeated. Immediately following the conclusion of each auditory stimulus, participants were asked to rate sound loudness and sound unpleasantness successively. One rating scale was used for each of the paradigms. The order of scale usage was pre-randomized for each participant. The mean of the two repetitions was calculated.

#### 2.4.4. Visual Stimuli

Participants completed four visual paradigms, similar to the visual paradigms used by Rosier and colleagues [24]. Participants were presented with eight different visual stimulus intensities in shades of gray (0, 14, 29, 44, 68, 72, 86, and 100% black), with each intensity printed in a 1 cm square in the center of an 8.5×11 white sheet of paper (one stimulus per sheet). Each stimulus intensity was presented separately and in a random order, such that in each paradigm, the participant would rate all eight stimulus intensities. The participant was asked to rate the darkness (or % black) of each stimulus as they were revealed. In each paradigm, the participants rated each stimulus intensity once, with only one of the rating scales (VAS or NRS). Thus, all intensities were rated four times by each participant (two times with each rating scale). The order of rating scale usage was pre-randomized for each participant. The mean of the repetitions was calculated.

### 2.5. Statistical Analysis

Analyses of different modalities involved slightly different numbers of participants where all ratings were successfully obtained: auditory intensity (n= 42), auditory unpleasantness (n= 35), heat intensity (n=46), heat unpleasantness (n= 46), and visual grey level intensity (n= 45). Four participants were excluded from the auditory analysis: three due to missing data (3 due to experimenter error, and one due to self-reported hearing difficulties). Auditory unpleasantness was not collected for 9 participants. Two participants were excluded from the visual analysis (one because the visual stimuli were not calibrated correctly and the other because one of the intensity levels was not tested).

Statistical analysis was conducted using M+ and R and Rstudio (version 3.6.2, Boston, MA, USA). Independent t-tests were used to identify sex differences between the groups with FDR corrections. In addition, separate ANOVA models were used for the heat, auditory and visual ratings. Greenhouse-Geiser correction was employed. The models assessed the differences between rating scales (VAS vs. NRS), stimulus intensities, sex and their interaction. Post hoc tests were used to identify specific differences. The models were conducted separately for pain intensity and for pain unpleasantness ratings. For the noxious heat model, there were 5 stimulus intensities for low heat (44-45), and 5 for high heat (47-48). For the auditory model, there were 4 stimulus intensities. For the visual model, there were 8 stimulus intensities of grey level.

The rating-based discrimination thresholds was also calculated to identify if one rating scale allows better discrimination between different stimulus intensities and if this is dependent on sex. Pain discrimination threshold was assessed for two reference temperatures (44°C and 48°C). Since pain sensitivity might affect discrimination to low or high temperatures, we performed these analyses at both the low end (44°C) and the high end (48°C) of the noxious heat range. 44°C-ascending discrimination thresholds were defined as the temperature closest to 44°C that had a 0.5 probability of being rated as more intense or unpleasant than the sensation at 44°C. Similarly, 48°C-descending discrimination thresholds were defined as the temperature closest to 48°C that had a 0.5 probability of being rated as less intense or unpleasant than the sensation at 48°C. The following steps were completed to define these thresholds: 1) individual ratings of the stimuli were binarized based on whether they were higher or lower than a reference rating, i.e. rating of 44°C stimuli for 44°C-ascending discrimination threshold, or the rating of 48°C stimuli for 48°C-descending discrimination threshold; 2) individual logistic regression was modeled, defining the individual β_0_ and β_1_ coefficients; 3) rating-based discrimination thresholds were calculated for a probability of 0.5 by dividing -β_0_ by β_1_. If participants reported changes in their pain intensity or unpleasantness compared to ratings of 44°C or 48°C in all the other trials, rating-based discrimination thresholds were defined at an arbitrary value that was closer to the reference than the minimal temperature change. This was to reflect that these participants were able to detect a change in temperature smaller than 0.2°C. Hence, 44°C-ascending discrimination thresholds were defined as 44.1°C and 48°C-descending discrimination thresholds were defined as 47.9°C.

The effect of ratings scale and sex on discrimination thresholds was examined. 2×2 ANOVA models for the 44°C-ascending and 48°C-descending discrimination thresholds were then conducted for scale, sex and their interaction. Because of violations of the assumption of normality, Kruskal-Wallis tests were performed. Data are presented as mean ± SD. A p-value of < 0.05 was considered significant.

## 3. Results

Forty-nine participants were enrolled. Of these, three participants were excluded from the study. One participant was taking a medication for a neurological disorder, and two participants tested positive for central nervous system-acting drugs on the urine drug screen. A total of forty-six participants with ages ranging from 14-41 years old completed the study (twenty-four women (ages 25.33±1.48), and twenty-two men (ages 25.09±1.67)). No differences between males and females were found for age or any of the psychological measures (Table 1).

**Table 1.** Sex differences in psychological measures.

| **Variable** | **Females**<br>(Mean ±SD) | **Males**<br>(Mean ±SD) | **t** | **df** | **p value** |
| --- | --- | --- | --- | --- | --- |
| **Age** | 25.3±7.2 | 25.1±7.8 | 0.109 | 44 | 0.914 |
| **Impulsiveness** | 53.3±7.8 | 58.5±9.5 | -2.012 | 44 | 0.050 |
| **Mindfulness** | 40.0±6.3 | 39.8 ±7.6 | 0.088 | 44 | 0.930 |
| **Positive affect** | 37.7±7.9 | 35.2±7.1 | 1.094 | 44 | 0.280 |
| **Negative affect** | 14.8±3.5 | 15.4±4.6 | -0.554 | 44 | 0.583 |
| **EPQ-R:** |  |  |  |  |  |
| **Neuroticism** | 4.0±2.7 | 3.1±2.7 | 1.126 | 43 | 0.266 |
| **Extraversion** | 8.3±3.3 | 7.2±3.2 | 1.116 | 43 | 0.270 |
| **Psychoticism** | 1.7±1.4 | 2.7±1.4 | -2.450 | 44 | 0.018 (not<br>significant after<br>FDR correction) |
| **Lie** | 6.7±3.1 | 5.2±3.4 | 1.536 | 43 | 0.132 |
| **Anxiety** | 12.4±6.2 | 11.8±6.8 | 0.290 | 44 | 0.773 |
| **Depression** | 8.8±3.6 | 9.7±4.8 | -0.714 | 44 | 0.479 |

EPQ-R: Eysenck Personality Questionnaire- revised edition

### 3.1 Heat stimuli

For both the low and high heat pain intensities, a significant effect for rating scale was found (p=0.001, Table 2, Fig 2) with NRS being rated higher compared to the VAS. Similarly, for both the low and high heat pain intensities, a significant effect for stimulus intensity was also found (p<0.001) indicating that higher temperatures were rated as more intense. The interaction between rating scale and stimulus intensity was not significant. In addition, all interactions with sex were not significant.

**Table 2.**
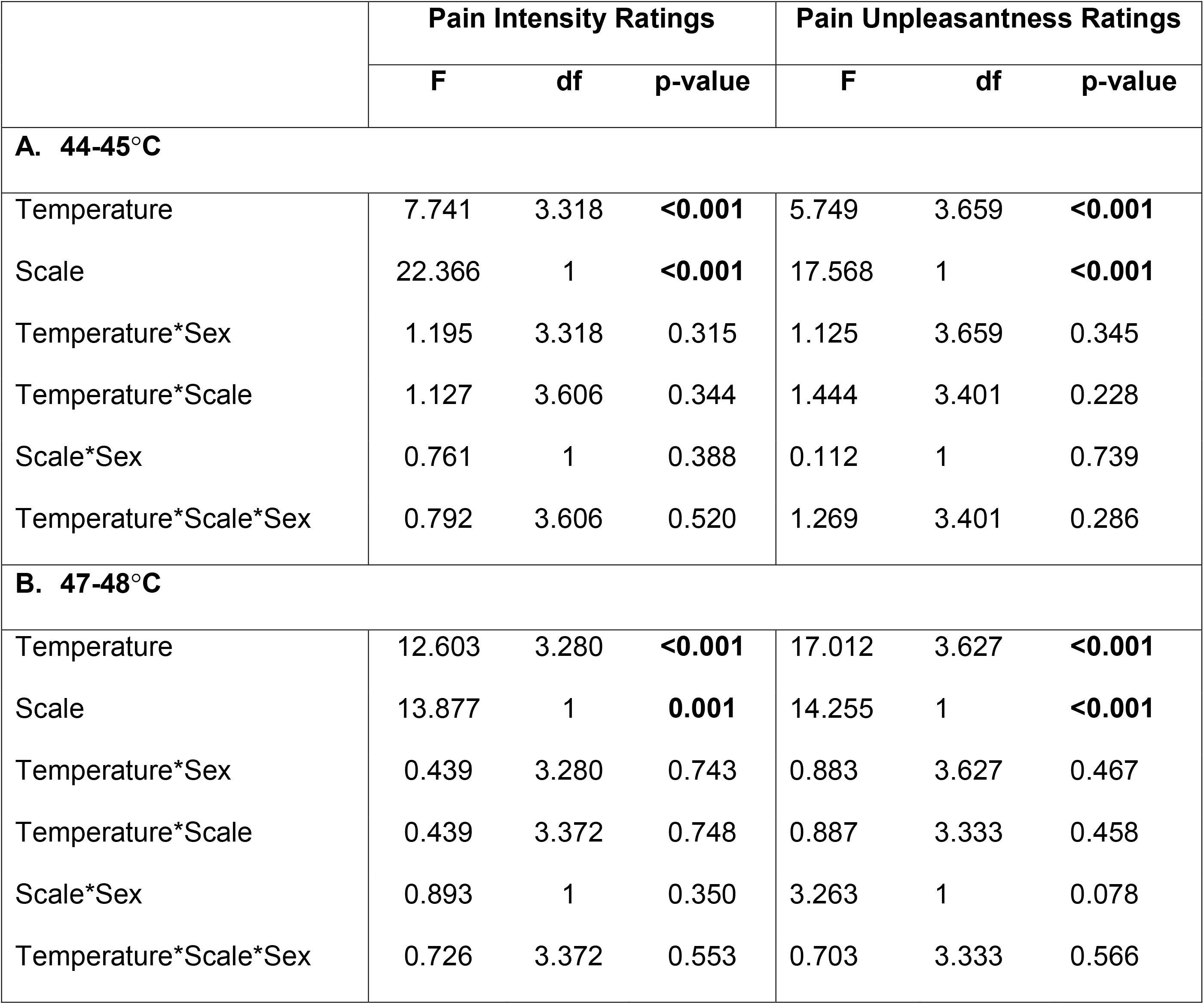
Pain Intensity and unpleasantness ratings.

|  | Pain Intensity Ratings |  |  | Pain Unpleasantness Ratings |  |  |
| --- | --- | --- | --- | --- | --- | --- |
|  | F | df | p-value | F | df | p-value |
| **A. 44-45°C** |  |  |  |  |  |  |
| Temperature | 7.741 | 3.318 | **<0.001** | 5.749 | 3.659 | **<0.001** |
| Scale | 22.366 | 1 | **<0.001** | 17.568 | 1 | **<0.001** |
| Temperature*Sex | 1.195 | 3.318 | 0.315 | 1.125 | 3.659 | 0.345 |
| Temperature*Scale | 1.127 | 3.606 | 0.344 | 1.444 | 3.401 | 0.228 |
| Scale*Sex | 0.761 | 1 | 0.388 | 0.112 | 1 | 0.739 |
| Temperature*Scale*Sex | 0.792 | 3.606 | 0.520 | 1.269 | 3.401 | 0.286 |
| **B. 47-48°C** |  |  |  |  |  |  |
| Temperature | 12.603 | 3.280 | **<0.001** | 17.012 | 3.627 | **<0.001** |
| Scale | 13.877 | 1 | **0.001** | 14.255 | 1 | **<0.001** |
| Temperature*Sex | 0.439 | 3.280 | 0.743 | 0.883 | 3.627 | 0.467 |
| Temperature*Scale | 0.439 | 3.372 | 0.748 | 0.887 | 3.333 | 0.458 |
| Scale*Sex | 0.893 | 1 | 0.350 | 3.263 | 1 | 0.078 |
| Temperature*Scale*Sex | 0.726 | 3.372 | 0.553 | 0.703 | 3.333 | 0.566 |

**Figure 2.**
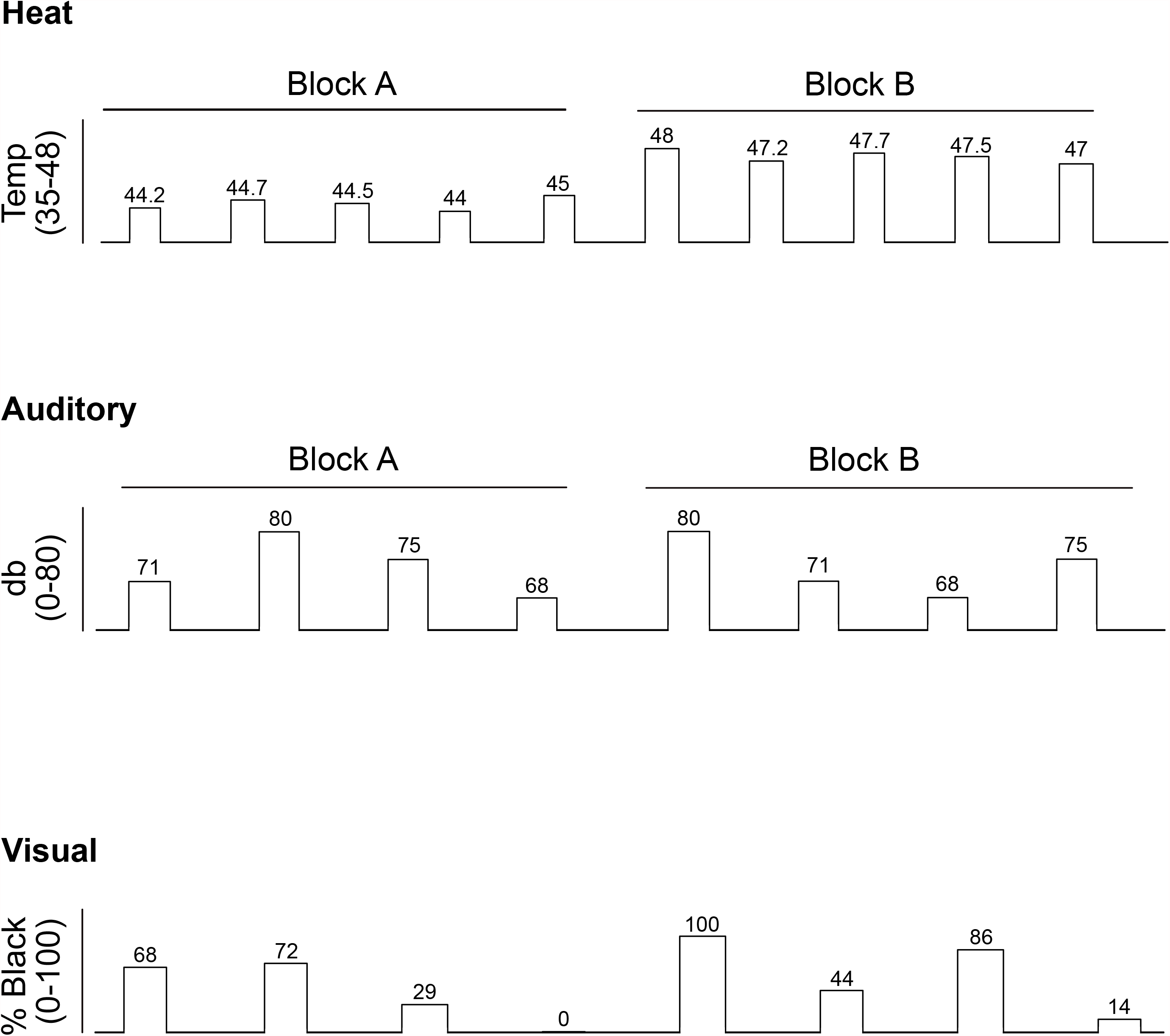
Pain ratings of noxious heat stimuli. Pain intensity and unpleasantness evoked by 44-45°C (A) and 47-48°C (B) stimuli were consistently rated higher using the NRS compared to the VAS. However, no interaction between scale and sex was identified.

Similar results were found for pain unpleasantness. A significant effect of rating scale was found for both the low and high heat pain intensities (p<0.001, Table 2, Fig 2). In addition, a significant effect for stimulus intensity was found (p<0.001). The interaction between rating scale and stimulus intensity was not significant. In addition, all interactions with sex were not significant.

#### 3.1.1. Discrimination thresholds to heat stimuli

The ability of participants to provide pain ratings that discriminated between small differences in temperatures was also examined. Ascending and descending pain discrimination thresholds were calculated. Both low and high reference temperatures were used (44°C for the ascending and 48°C for the descending discrimination threshold). 44°C-ascending discrimination thresholds were defined as the temperature closest to 44°C that had a 0.5 probability of being rated as more intense or unpleasant than the sensation at 44°C. Similarly, 48°C-descending discrimination thresholds were defined as the temperature closest to 48°C that had a 0.5 probability of being rated as less intense or unpleasant than the sensation at 48°C.

A trend for scale was observed for discrimination thresholds of pain intensity (p=0.077) and pain unpleasantness (p=0.065) ratings, with lower discrimination thresholds for the VAS. For pain intensity, in the ascending discrimination thresholds, males and females were able to discriminate between temperature differences of 0.71 and 0.73°C, respectively with the VAS compared to 0.73 and 0.79°C with the NRS. For pain intensity, in the descending discrimination thresholds, males and females were able to discriminate between temperature differences of 0.52 and 0.45°C, respectively with the VAS compared to 0.73 and 0.71°C with the NRS. No effect for scale, sex or their interaction on discrimination thresholds (ascending and descending) was observed. This was found for both pain intensity ratings and pain unpleasantness ratings (Table 3, Fig. 3).

**Table 3.**
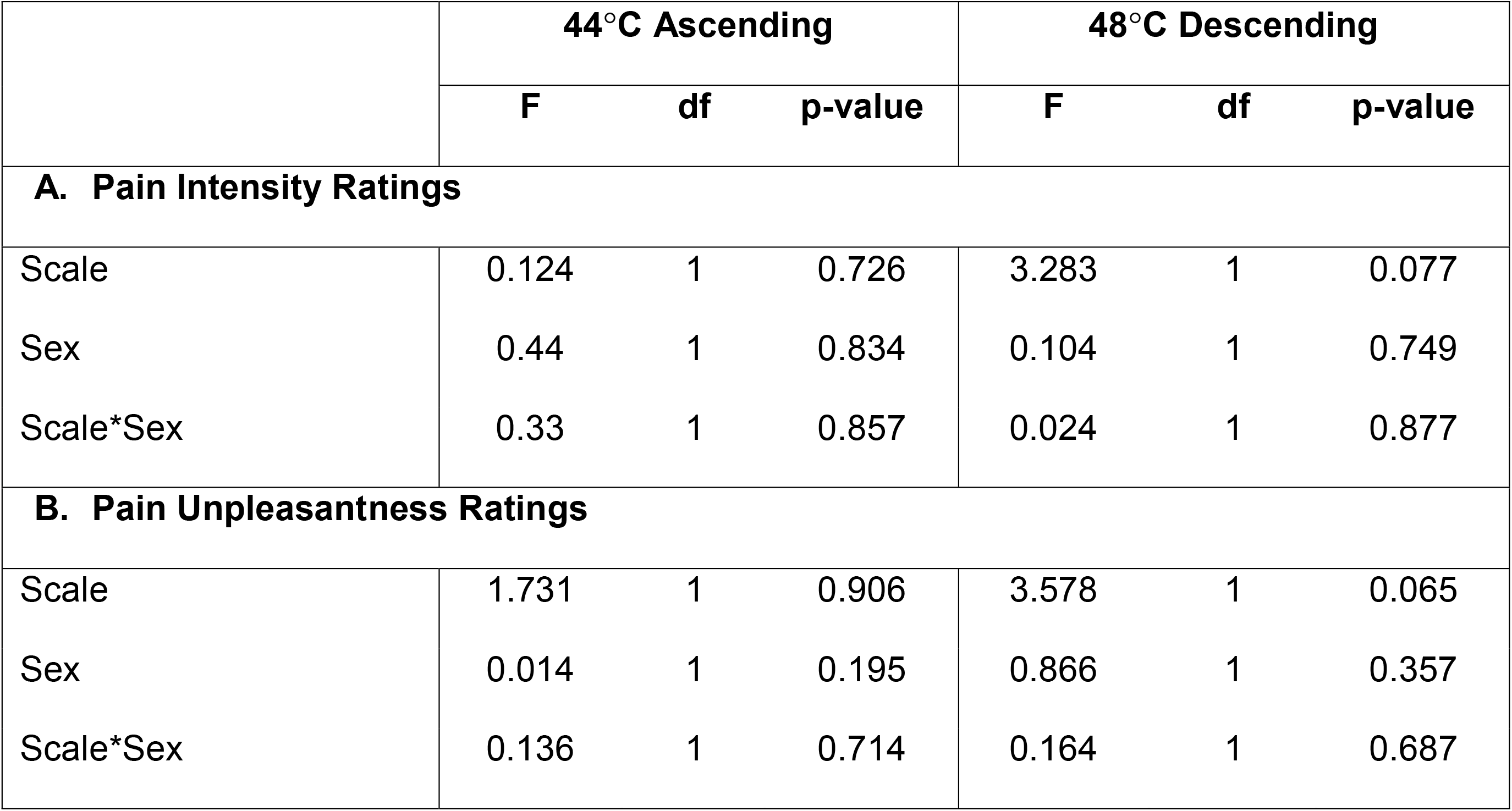
Discrimination threshold.

**Figure 3.**
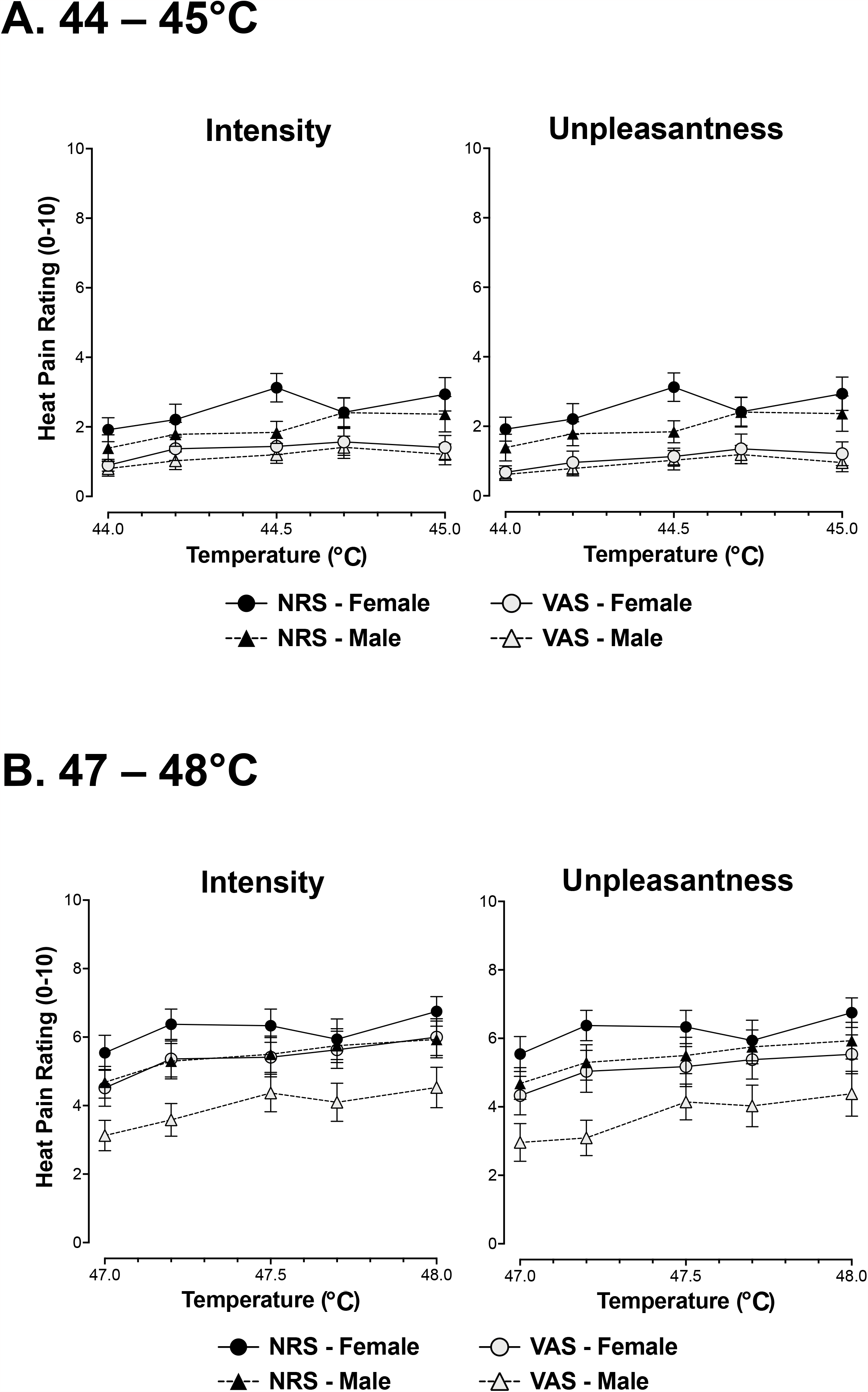
Discrimination thresholds to heat stimuli. **A**. Ascending discrimination thresholds were calculated as the minimal temperature difference that a participant rated differently starting from a reference temperature of 44°C. A trend towards an effect of scale was noted, but no interactions between scales and no interactions between rating scale and sex were found for pain intensity and unpleasantness. **B**. Descending discrimination thresholds were calculated as the minimal temperature difference that a participant rated differently starting from a reference temperature of 48°C. A trend towards an effect of scale was noted, but no interactions between rating scale and sex were found for pain intensity and unpleasantness.

### 3.2 Auditory stimuli

For sound intensity, a significant effect for rating scale was found (p<0.001, Table 4, Fig. 4) with stimulus intensity being rated higher with NRS compared to the VAS. A significant effect for sound level was also found (p<0.001). The interaction between rating scale and sound level was not significant. In addition, all interactions with sex were not significant.

**Table 4.** Auditory Intensity and unpleasantness ratings.

|  | **F** | **df** | **p-value** |
| --- | --- | --- | --- |
| **A. Auditory Intensity Ratings** |  |  |  |
| Sound Level | 142.030 | 3 | **<0.001** |
| Scale | 44.065 | 1 | **<0.001** |
| Sound Level *Sex | 0.317 | 1.567 | 0.676 |
| Sound Level *Scale | 1.253 | 1.939 | 0.291 |
| Scale*Sex | 0.001 | 1 | 0.975 |
| Sound Level *Scale*Sex | 1.625 | 1.939 | 0.204 |
| **B. Auditory Unpleasantness Ratings** |  |  |  |
| Sound Level | 101.471 | 1.842 | **<0.001** |
| Scale | 78.564 | 1 | **<0.001** |
| Sound Level *Sex | 0.706 | 1.842 | 0.487 |
| Sound Level *Scale | 4.154 | 1.756 | **0.025** |
| Scale*Sex | 0.549 | 1 | 0.464 |
| Sound Level *Scale*Sex | 2.382 | 1.756 | 0.108 |

**Figure 4.**
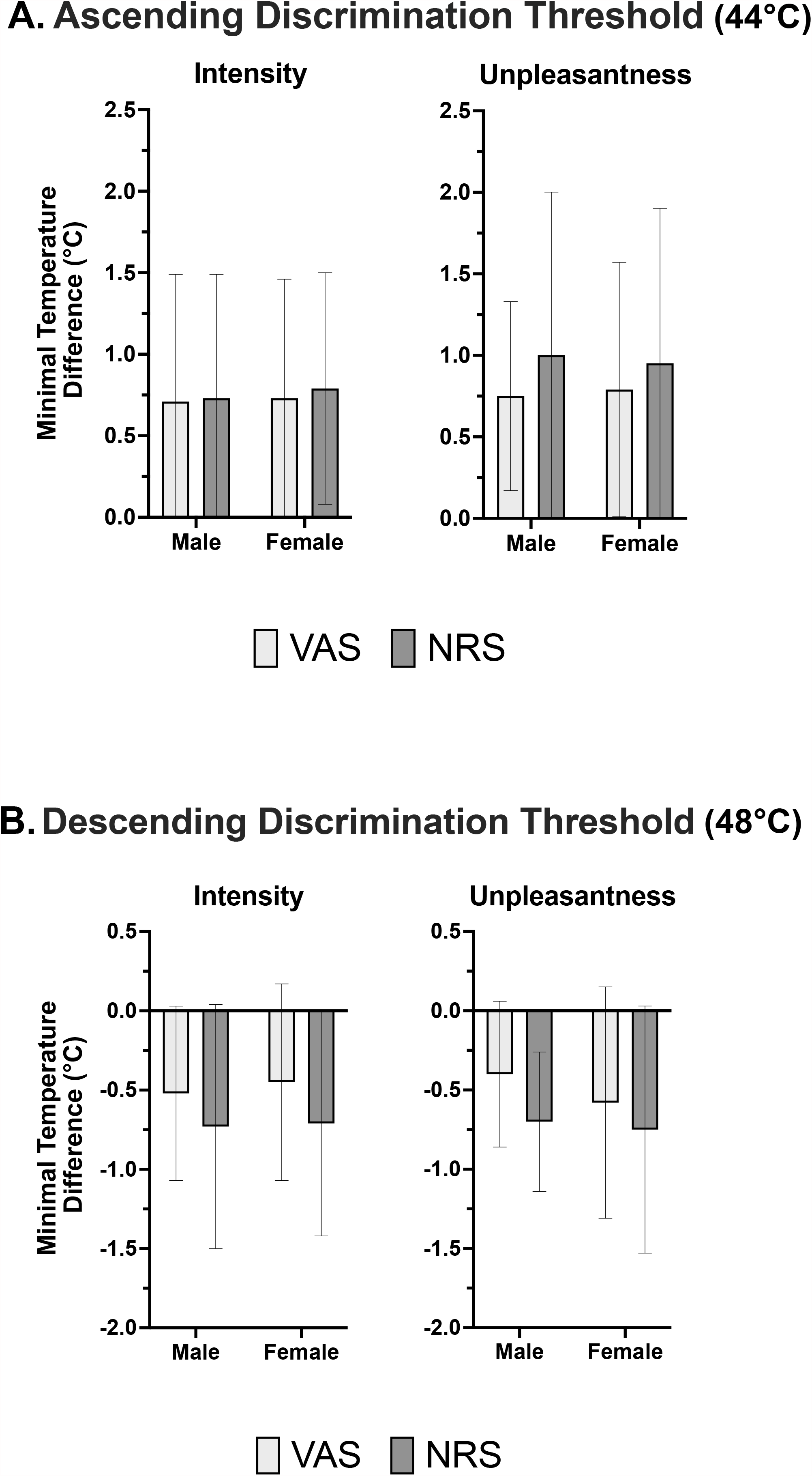
Intensity and unpleasantness ratings of auditory stimuli. Intensity and unpleasantness ratings of auditory stimuli were rated higher using the NRS compared to the VAS. No interactions between rating scale and sex were found for intensity and unpleasantness.

Similar results were found for sound unpleasantness. A significant effect for rating scale was found (p<0.001, Table 4, Fig. 4), as well as a significant effect for sound level (p<0.001). The interaction between rating scale and sound level was also significant (p=0.025) indicating that for all sound levels, sound unpleasantness was rated higher with the NPS compared to VAS. In addition, the sex X rating scale interaction, sex X sound unpleasantness rating interaction, and sex X rating scale X sound unpleasantness rating interaction were not significant.

### 3.3 Visual stimuli

For visual stimuli, a significant effect for rating scale was found (p<0.001, Table 5, Fig. 5) with grey level (% black) being rated higher with the NRS compared to the VAS. A significant effect for grey level was also found (p<0.001). The interaction between rating scale and grey level was significant (p<0.001) with all levels being rated significantly higher with the NRS compared to the VAS, except for the 0% and 100% black in which no significant differences in ratings scales were found. All interactions with sex were not significant.

**Table 5.** Visual ratings of grey level (% black)

|  | **F** | **df** | **p-value** |
| --- | --- | --- | --- |
| Grey level | 1038.704 | 3.342 | **<0.001** |
| Scale | 38.537 | 1 | **<0.001** |
| Grey level *Sex | 0.587 | 3.342 | 0.643 |
| Grey level *Scale | 5.901 | 5.028 | **<0.001** |
| Scale*Sex | 0.116 | 1 | 0.735 |
| Grey level *Scale*Sex | 0.370 | 5.028 | 0.870 |

**Figure 5.**
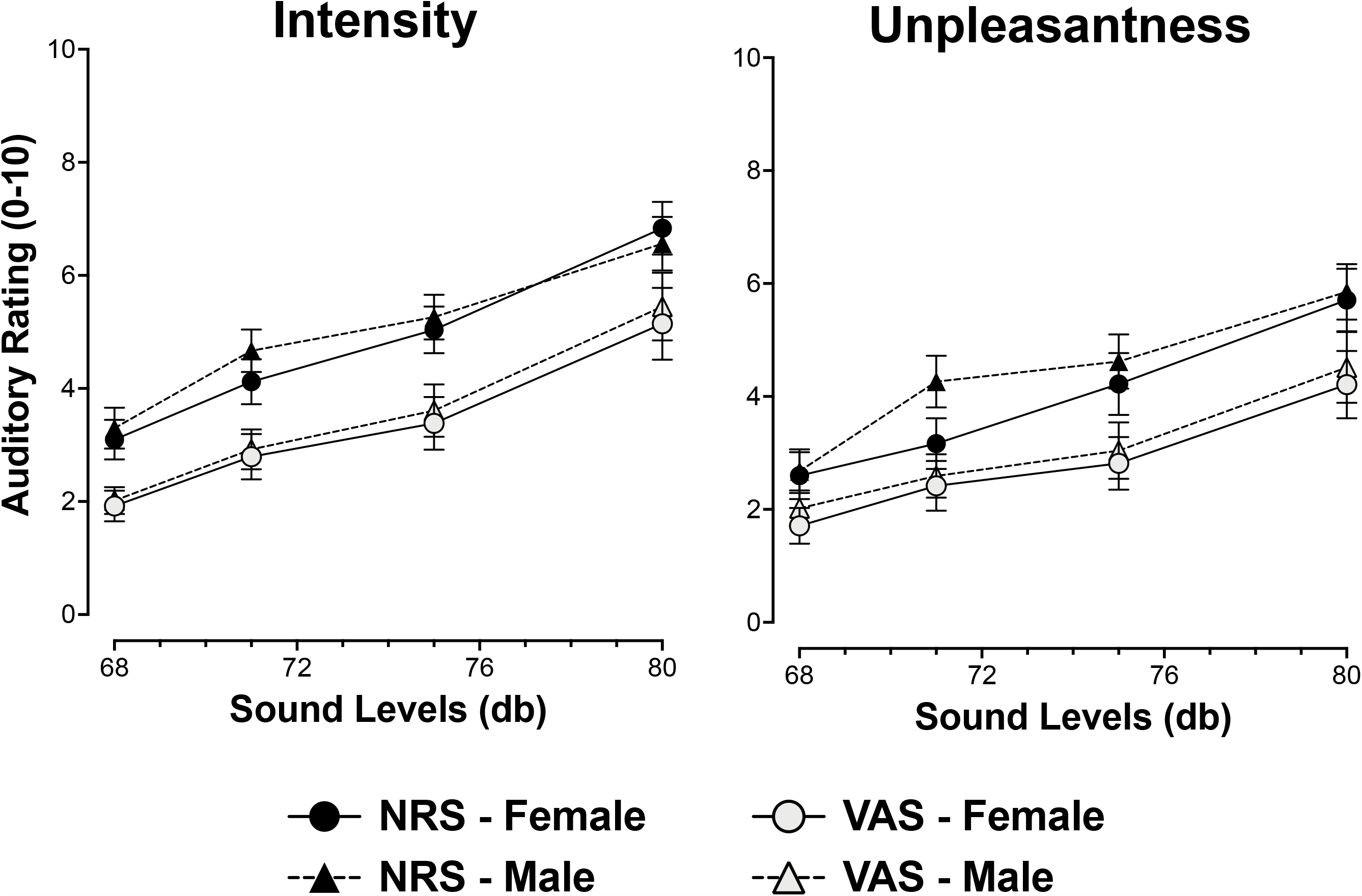
Ratings of visual stimuli. Intensity ratings of visual stimuli (% black) were rated higher using the NRS compared to the VAS. No interactions between rating scale and sex were found for intensity.

## 4. Discussion

Across all modalities (noxious heat, auditory and visual) participants provided higher ratings for the same stimulus intensities when they were using the NRS compared to VAS. Different usage of rating scales between males and females was not found, suggesting that biases in rating scale usage may contribute negligibly to sex differences in pain. Thus, there is a bias between rating scales, but it is similar across participants and across modalities.

### 4.1 Differences between the NRS and VAS

Ratings obtained with VAS and NRS are typically highly correlated [2, 3, 8, 10, 11, 14, 26]. However, key differences between them exist. The VAS has been shown to have linear, ratio scale properties [16, 20]. For example, healthy participants and patients with chronic pain completed a stimulus-response task in which they rated pain intensity and pain unpleasantness. Participants rated 6 noxious heat stimuli ranging from 43-51°C. Then, participants were asked to select a temperature that is perceived as twice as painful as the pain evoked by a 43°C and 45°C temperature. In addition, pairs of stimuli were presented, and participants were asked to indicate the ratio of how much more painful is the high intensity stimulus compared to the low intensity stimulus. The stimulus response-function was used to predict both ratio and the temperature that is as twice as intense as the 43°C or 45°C temperature. These results indicated that the VAS has a ratio scale property [20]. A later similar study from the same group examined the ratio scale of the NRS in comparison to the VAS. While the ratio scaling for the VAS were again found, the NRS however, did not have ratio properties [19].

In contrast, even though NRS is considered to be a continuous measure, participants may use it as a categorical measure [6]. When using a 0-100 NRS, participants are most likely to use integers of 10 or 5, despite the fact that there are options to use other integers [12]. However, when rating pain with the VAS, participants are more likely to use the whole scale, which may make the VAS more sensitive to small changes in pain perception [12]. The use of integers for the NRS may be a function of instruction. For clinical purposes, a whole integer scale may be adequate – individual patient improvements in fractions of a point are meaningless. However, in aggregate groups of patients and/or studies using carefully controlled stimuli, such potential differences in sensitivity may be significant. So, there may be stronger implications for differences between NRS and VAS for research than in routine clinical use. In the present study, only three participants used decimals for the NRS. Thus, a sub-analysis comparing NRS ratings of only integers vs. of decimals was not possible.

### 4.2 Participants provide higher ratings with the NRS

In the present investigation, higher ratings were consistently found with the NRS. Thus, even though prior findings support the notion of a strong correlation between scales, correlations do not take consistent shifts in the magnitude of ratings with one scale into account. The finding of higher ratings with the NRS is in agreement with previous studies, including studies in adult pain patients who rated noxious heat stimuli [3, 11, 19], and a study with pediatric patients with sickle cell disease, who rated their clinical pain [17].

Higher ratings with the NRS were found for all the tested modalities (noxious heat, auditory and visual). This suggests that ratings obtained with the NRS are subject to a bias related to the scale usage rather than to the evaluation of specific stimuli of certain modalities. Price and colleagues have suggested that the higher pain ratings observed with the NRS could be due to number perseverance. In this bias, participants consistently choose a specific number and use it to rate different intensities of stimuli. Another suggestion was that with the NRS, participants choose to assign different numbers to different stimuli [19].

### 4.3 Discrimination thresholds

The discrimination thresholds were examined to address differences between the rating scales, and specifically, the suggestion that when using the NRS, participants may persevere with a given number [19]. Based on this, we predicted that the participants would be able to discriminate between smaller stimulus intensity differences when using the VAS compared to the NRS (i.e. participants would have a smaller discrimination threshold when using the VAS vs. the NRS). A trend for scale was observed for discrimination thresholds of pain intensity and pain unpleasantness ratings, with lower discrimination thresholds for the VAS. However, no sex effect or interaction between sex and scale were found for discrimination thresholds, indicating that both scales allow male and female participants to detect small differences in temperature. A previous study found that ratings obtained from participants using the VAS can better discriminate between pairs of adjacent heat stimuli (difference of 1°C) compared to a verbal descriptor scale [24]. In addition, NRSs of 11, 21 and 101 points are better in detecting small changes in pain perception compared to NRSs of <6 points [12]. Another study found no differences between rating scales in distinguishing between 0.5°C increments [21]. The present study did not find differences between the VAS and NRS rating scales in discrimination thresholds using smaller differences of 0.2-0.3°C. This suggests that both scales are sufficiently sensitive to detect small temperature changes.

### 4.4 Sex differences in scale usage

Sex differences in experimental pain have been extensively investigated, and conflicting results have been found [4, 15, 22, 23]. Different usage of scales by males and females is one potential confound when examining sex differences in pain in that each sex could experience a similar sensation, but report it differently (or vice-versa) [21]. A previous study compared pain ratings to noxious heat using VAS, a number box numerical scale, which involves sliding a track ball to get to the desirable number, and a combined VAS with a number box numerical scale. Sex differences were found for scale usage such that healthy females had greater pain ratings compared to males when using the number box numerical scale. However, this was not found for the other VAS scales [21]. The method of number box numerical scale is very different from the more common method of NRS. In both cases participants are asked to rate a sensation using a number. However, in the NRS method, participants give a verbal answer or mark their response on a paper, while in the number box numerical scale participants scroll through all the numbers until they reach the desired rating score. In another study examining only cold pain, sex differences in pain ratings were found for NRS, verbal rating scale and faces pain scales. However, sex differences were not found when participants used the VAS [8]. Taken together, these observations raise the possibility that sex differences in rating scale usage may confound the assessment of sex differences in pain and that studies using NRS may be biased to find positive sex differences in pain. In contrast to these possibilities, the present study found no differences between females and males for both NRS and VAS for noxious heat, auditory and visual stimuli.

Minimal sex biases in scale usage have been reported when multiple scales were used to assess chronic pain [25]. Patients with chronic pain rated their pain using a VAS, verbal rating scale (pain descriptors) and faces pain scale. Sex was not a predictor for pain ratings for any of the scales, and explained only 3% or less of the variability in pain ratings [25].

### 4.5 Study strengths and limitations

A strength of this study is the inclusion of various stimulus modalities (noxious heat, auditory and visual). In addition, the study design randomized between paradigms and stimuli within paradigms. Thus, it was possible to differentiate between changes in pain sensitivity and scale effect. Accordingly, we determined that the increased pain ratings found with NRS compared to VAS across all modalities, represent biases of scale usage.

A limitation of the study is that we did not examine the participants’ scale preference. Participants may find the usage of VAS more challenging and prefer the NRS or vice versa. However, the present study included an extensive familiarization component (16 stimuli) in which participants experienced and were familiarized with using both rating scales. Mimicking clinical practice, participants were not explicitly instructed to use fractions; however, this may influence the level of discrimination. A previous study that compared three pain scales, found that after completing a similar familiarization part, participants reported that they are comfortable with using the pain scales [21]. This should minimize error usage of rating scales and ensure proper usage in both scales, even if participants prefer one over the other.

## 5. Conclusion

Sex differences in pain sensitivity are probably not due to different usage of the NRS or VAS rating scales. There is an overall bias in which ratings for auditory, visual and somatosensory stimuli are higher with NRS compared to VAS. Choosing a rating scale for research or clinical use should take this bias into account.

## Data Availability

Data will be shared upon request.

## Acknowledgements

We wish to thank Dr. Kenneth R Goldschneider for his insightful comments on the manuscript. This study was funded by a NIH/NINDS (R01NS085391, RCC) grant.

---

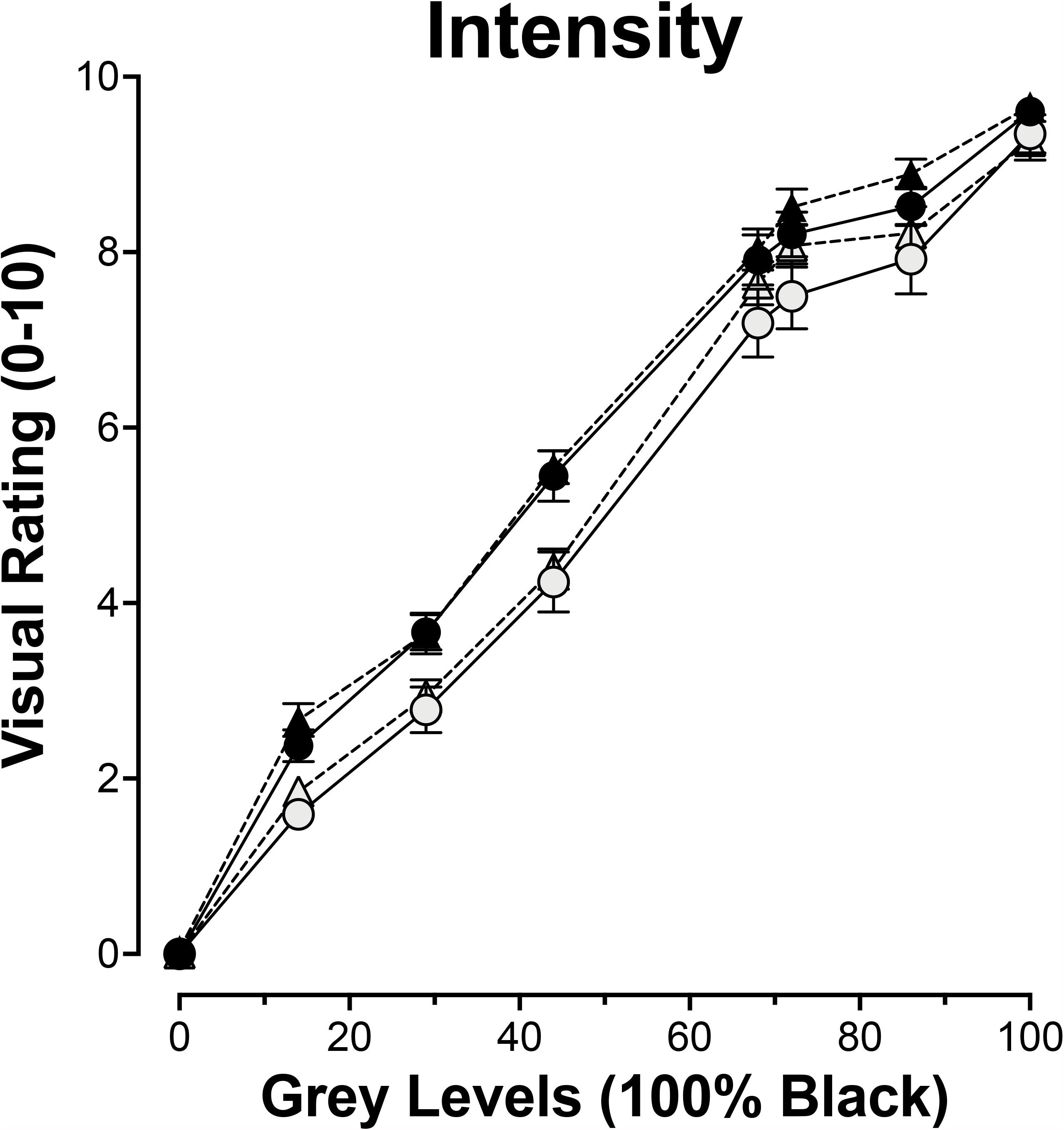

